# Elevated renin activity and change in sleep apnea with primary aldosteronism

**DOI:** 10.1101/2024.08.01.24311390

**Authors:** Yonekazu Kidawara, Manabu Kadoya, Masataka Igeta, Miki Kakutani-Hatayama, Akiko Morimoto, Akio Miyoshi, Akinori Kanzaki, Kosuke Konishi, Takashi Daimon, Hidenori Koyama

**Author notes:** Corresponding author: Manabu Kadoya, M.D., Ph.D. Department of Diabetes, Endocrinology and Clinical Immunology, School of Medicine, Hyogo Medical University 1-1 Mukogawa-cho, Nishinomiya, Hyogo 663-8501, Japan.

## Abstract

**Background:** The prevalence of obstructive sleep apnea (OSA) is higher in patients with primary aldosteronism (PA), while elevated renin activity after treatment is associated with a lower risk of cardiovascular events. However, the association of PA with degree of OSA remains unclear and it is not known whether elevated renin activity in PA patients is associated with change in apnea condition.

**Methods:** Cross-sectional relationships between PA (n=176) and degree of OSA classified by apnea-hypopnea index (AHI) with use of an apnomonitor were investigated, with the results compared with those obtained with non-PA patients (n=418). Additionally, the effects of elevated renin activity on change in AHI were prospectively examined in 45 patients with PA.

**Results:** Patients with PA were found to be significantly associated with severe OSA even after adjustment for other clinical risk factors (odds ratio 2.08, 95% confidence interval 1.09-3.95, p = 0.025) as compared to those without PA. Furthermore, the logarithm of the renin activity after treatment and change in AHI before and after treatment were significantly negatively correlated, with Pearson’s correlation coefficient (r = -0.364, p = 0.014).

**Conclusions:** Severe OSA is more commonly seen in PA patients with hypertension as compared to patients without PA, and elevated renin activity may contribute to improvement of sleep apnea in patients with PA.

## Introduction

Primary aldosteronism (PA) is recognized as one of the most common causes of secondary hypertension, and affected individuals have also been shown to have a higher risk for cardiovascular events and mortality independent of blood pressure level as compared to patients with essential hypertension (EH)^1–4^. Obstructive sleep apnea (OSA), another well-known cause of hypertension, has also been found to contribute to increased risk of cardiovascular morbidity and mortality ^5–7^. In patients with PA, the prevalence of OSA has been reported to be as high as 60% to 80% ^8^. Additionally, those patients have a tendency for higher OSA prevalence as compared to patients without PA who are hypertension resistant ^9^. Another report regarding risk of PA showed that individuals with maintained suppression of renin activity had increased risk of cardiovascular events and death ^10^. Therefore, unsuppressed renin activity is now recognized as a therapeutic goal for PA patients, though it remains unclear whether elevated renin activity is associated with changes related to apnea. The present study was performed as a cross-sectional examination of the association of PA with degree of OSA and also prospectively investigated the effect of elevated renin activity on sleep apnea.

## Methods

### Study design and participants

From October 2010 to November 2020, 1151 patients assessed for diagnosis of PA and OSA were enrolled in the Hyogo Sleep Cardio-Autonomic Atherosclerosis (HSCAA) study. Out of these, 781 were hypertensive patients including those with PA. For the present study, those with autonomous cortisol secretion, pheochromocytoma, primary hyperparathyroidism, estimated glomerular filtration rate (eGFR) <15 mL/min/1.73 m^2^, or dialysis treatment were excluded (n=128). Additionally, 59 with missing baseline data regarding assessment of OSA were also excluded. Finally, 594 were enrolled, and divided into the PA (n=176) and non-PA (n=418) groups. Of those with PA, 45 patients were simultaneously reassessment for renin activity and sleep apnea within five years (**Figure 1**). HSCAA study, which has received approved by the institutional ethical committee of Hyogo Medical University (approval No. 2351), was used and informed written consent was obtained from each participant.

**Figure 1.**
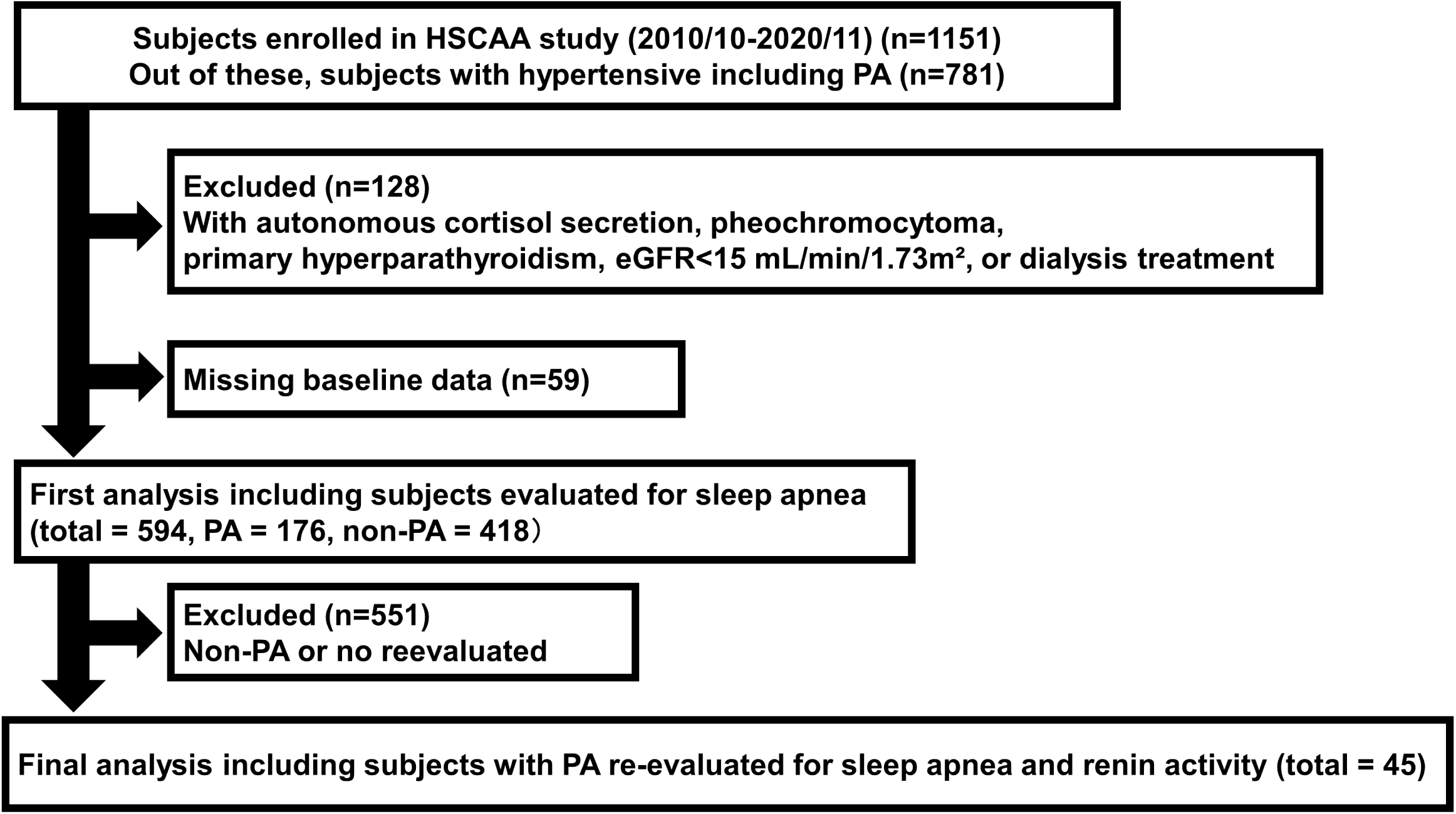
Flow of subject selection. eGFR, estimated glomerular filtration rate; PA, primary aldosteronis.

### Plasma biochemical parameters

Blood samples were obtained in the morning after an overnight fast and then quickly centrifuged to obtain plasma. Whole blood was used for hemoglobin A1c, EDTA plasma for glucose and lipids, and serum for other biochemical assays. Glucose measurements were performed using a glucose oxidase method and an enzymatic method was used to determine serum creatinine concentration. Estimated glomerular filtration rate in each patient was calculated using an equation for Japanese subjects, as follows: estimated glomerular filtration rate (mL/min per 1.73 m^2^)=194×age (years)^-0.287^×S-creatinine^-1.094^ (if female,×0.739). Plasma renin activity (PRA) and plasma aldosterone concentration (PAC) were measured using an enzyme immunoassay with the patient at rest in a supine position for 30 minutes.

### Diagnosis of primary aldosteronism

Patients were diagnosed with PA based on a PAC (pg/mL) to PRA (ng/mL/hour) ratio (ARR) ≥200 at screening and at least one positive result from confirmatory tests for PA diagnosis (saline infusion test, captopril challenge test, furosemide-upright test) at the time of admission, according to previously reported guidelines ^11,12^.

### Assessment of sleep apnea

To determine the presence of sleep apnea, an apnomonitor device (SAS-2100, Nihon Kohden, Tokyo, Japan; supplied by Teijin, Tokyo, Japan) was used to provide apnea-hypopnea index (AHI) values. Criteria presented in The American Academy of Sleep Medicine Manual for the Scoring of Sleep and Associated Events, version 2.1, were used for analysis of respiratory events, as follows.

Apnea was defined when the amplitude ratio to normal breathing was ≤10% with air flow lasting ≥10 seconds, and hypopnea when the amplitude ratio to normal breathing was ≤70% with air flow lasting ≥10 seconds and associated with a 4% decrease in oxygen saturation. AHI was calculated as the average number of apnea and hypopnea episodes per hour. When the decrease was 3% or more per second or the sensor was disconnected, the result was considered an artifact and excluded from analysis. The degree of sleep apnea was classified based on AHI values as none (0 to <5), mild (5 to <15), moderate (15 to <30), or severe (≥30) ^13^.

### Statistical analyses

For comparisons of mean and median values of continuous variables for the baseline clinical characteristics between PA status (PA and non-PA), and also suppressed renin activity status in patients with PA (remained suppressed and unsuppressed), Welch’s t-test or the Mann-Whitney U-test was used. Fisher’s exact test was used for categorical variables. Multivariable logistic regression analysis was used to calculate odds ratio (OR) and 95% confidence interval (CI) values for the associations of degree of OSA with PA as compared to the non-PA group adjusting for clinical factors, including potential causal factors for sleep apnea, i.e., age, male gender, BMI, systolic BP, HbA1c, and renal function. In addition, a Pearson’s correlation coefficient was calculated to evaluate the association between the change of AHI and elevation of the renin activity in PA patients. The logarithmic transformation was performed to the renin activity for a normal approximation. All statistical analyses were performed with EZR, version 1.61 (Saitama Medical Center, Jichi Medical University, Saitama, Japan) and SAS software version 9.4 (SAS Institute Inc., Cary, NC, USA). All reported P values are two-sided and considered to be statistically significant at <0.05 without adjustment for multiplicity.

## Results

### Baseline clinical characteristics

The baseline characteristics of the present patients after dividing into the PA and non-PA groups are shown in **Table 1**. The distribution of degree of sleep apnea in those with PA was as follows: normal (AHI <5), 26.7%; mild (AHI 5 to <15), 38.6%; moderate (AHI 15 to <30), 18.8%; and severe (AHI ≥30), 15.9%. Patients with PA exhibited higher levels of systolic BP, diastolic BP, and plasma aldosterone concentration, lower levels of fasting plasma glucose, HbA1c, and plasma renin activity, and lower prevalence of diabetes and dyslipidemia, as compared to the non-PA group.

**Table 1.** Comparisons of baseline clinical characteristics

| Variables | PA | Non-PA | P |
| --- | --- | --- | --- |
| Number of patients | 176 | 418 |  |
| Age, years | 57.1 ± 12.8 | 63.7 ± 12.2 | <0.001 |
| Male, n (%) | 84 (47.7) | 231 (55.3) | 0.112 |
| Body mass index, kg/m<sup>2</sup> | 25.4 ± 4.9 | 25.3 ± 5.2 | 0.682 |
| Current smoker, n (%) | 41 (23.3) | 113 (27.0) | 0.397 |
| Systolic BP, mmHg | 137.2 ± 15.7 | 131.2 ± 14.4 | <0.001 |
| Diastolic BP, mmHg | 82.4 ± 10.3 | 76.4 ± 9.0 | <0.001 |
| LDL cholesterol, mg/dL | 112.6 ± 29.4 | 111.8 ± 32.5 | 0.818 |
| HDL cholesterol, mg/dL | 54.8 ± 15.9 | 53.1 ± 17.0 | 0.273 |
| Triacylglycerol, mg/dL | 120.5 (84.0 – 165.3) | 115.0 (85.0 – 171.0) | 0.911 |
| eGFR, mL/min/1.73 m<sup>2</sup> | 80.0 ± 19.2 | 77.6 ± 23.0 | 0.224 |
| Fasting plasma glucose, mg/dL | 98 (91 - 106) | 101 (93 - 121) | <0.001 |
| HbA1c (%) | 5.5 (5.2 - 5.9) | 5.9 (5.3 - 6.8) | <0.001 |
| Plasma renin activity, ng/mL/hour | 0.4 (0.2 - 0.6) | 0.9 (0.5 - 2.1) | <0.001 |
| Plasma aldosterone concentration, pg/mL | 212.5 (153.0 – 293.8) | 102.0 (63.6 – 150.0) | <0.001 |
| Comorbidities |  |  |  |
| Diabetes, n (%) | 29 (16.5) | 197 (47.1) | <0.001 |
| Dyslipidemia, n (%) | 99 (56.3) | 284 (67.9) | 0.009 |
| Anti-hypertensive drugs |  |  |  |
| Calcium channel blocker, n (%) | 108 (71.5) | 262 (68.8) | 0.604 |
| α blocker, n (%) | 12 (7.9) | 31 (8.1) | 1.000 |
| Diuretic agent, n (%) | 18 (11.9) | 34 (8.9) | 0.375 |
| Sleep apnea |  |  |  |
| AHI, n (%) | 10.0 (4.8-20.3) | 9.7 (3.9-19.6) | 0.281 |
| Normal (<5), n (%) | 47 (26.7) | 125 (29.9) | 0.493 |
| Mild (5 to <15), n (%) | 68 (38.6) | 145 (34.7) | 0.411 |
| Moderate (15 to <30), n (%) | 33 (18.8) | 104 (24.9) | 0.130 |
| Severe (AHI ≥30), n (%) | 28 (15.9) | 44 (10.5) | 0.090 |

Data are presented as the mean ± standard deviation or median (25th-75th percentile) for continuous variables or number (%) for categorical dichotomous variables. P values based on the two-sample Welch's t-test or Mann-Whitney U-test are shown for comparisons of location parameters, such as the mean or median of two groups, or P values based on the Fisher's exact test for comparisons of percentage between groups. PA, primary aldosteronism; BP, blood pressure; LDL, low density lipoprotein; HDL, high density lipoprotein; eGFR, estimated glomerular filtration rate; AHI, apnea-hypopnea index

The baseline clinical characteristics of the patients classified by degree of OSA are presented in **Table 2**. Comparisons were also made between patients classified according to degree of OSA and those without OSA. There were no clinically significant differences in regard to smoking, diastolic BP, LDL-cholesterol, PRA, PAC, or use of anti-hypertensive drugs such as calcium-channel blocker and diuretic agent between patients with OSA regardless of severity as compared to those without OSA. Patients with OSA showed higher levels of BMI, systolic BP, FPG, and HbA1c, and a higher prevalence for male gender and diabetes, and lower level of eGFR as compared to patients without OSA. Those with mild or moderate OSA were older than those without OSA. As compared to patients without OSA, those with moderate or severe OSA had lower levels of HDL, higher levels of TG, and a higher prevalence of dyslipidemia. Users of α blocker anti-hypertensive drugs were more frequently noted among patients with severe OSA as compared to those without OSA. Furthermore, the prevalence of PA tended to be higher in patients with severe OSA, though the difference was not significant as compared to patients classified with other levels of OSA.

**Table 2.** Comparisons of baseline clinical characteristics among subjects divided based on degree of sleep apnea

| Variables | Normal<br>(AHI <5) | Mild<br>(AHI 5 to <15) | Moderate<br>(AHI 15 to <30) | Severe<br>(AHI ≥30) | P value<br>(Normal vs.<br>Mild) | P value<br>(Normal vs.<br>Moderate) | P value<br>(Normal vs.<br>Severe) |
| --- | --- | --- | --- | --- | --- | --- | --- |
| Number | 170 | 214 | 138 | 72 | - | - | - |
| Age, years | 59.3 ± 14.0 | 62.2 ± 11.7 | 64.9 ± 11.4 | 60.6 ± 13.8 | 0.026 | <0.001 | 0.489 |
| Male, n (%) | 74 (43.5) | 117 (54.7) | 81 (58.7) | 43 (59.7) | 0.039 | 0.011 | 0.031 |
| Body mass index, kg/m<sup>2</sup> | 22.9 ± 3.6 | 25.1 ± 4.7 | 26.1 ± 4.8 | 30.1 ± 6.4 | <0.001 | <0.001 | <0.001 |
| Current smoker, n (%) | 37 (21.8) | 59 (27.6) | 34 (24.6) | 24 (33.3) | 0.235 | 0.646 | 0.083 |
| Systolic BP, mmHg | 130.2 ± 14.5 | 133.6 ± 15.3 | 133.6 ± 15.3 | 136.4 ± 14.3 | 0.024 | 0.045 | 0.002 |
| Diastolic BP, mmHg | 77.6 ± 10.1 | 78.9 ± 9.6 | 77.6 ± 9.5 | 78.7 ± 10.2 | 0.221 | 0.990 | 0.458 |
| LDL cholesterol, mg/dL | 112.8 ± 33.4 | 110.0 ± 31.5 | 113.2 ± 28.8 | 114.2 ± 32.5 | 0.540 | 0.930 | 0.798 |
| HDL cholesterol, mg/dL | 55.5 ± 18.0 | 54.9 ± 16.0 | 51.0 ± 15.1 | 50.3 ± 17.3 | 0.709 | 0.021 | 0.039 |
| Triacylglycerol, mg/dL | 102 (78 - 147) | 115 (84 - 154) | 125 (96 - 179) | 141 (106 - 194) | 0.203 | 0.002 | <0.001 |
| eGFR, mL/min/1.73m<sup>2</sup> | 83.0 ± 21.9 | 78.5 ± 21.8 | 75.6 ± 20.4 | 72.1 ± 23.6 | 0.043 | 0.002 | <0.001 |
| Fasting plasma glucose, mg/dL | 97 (90 - 105) | 101 (93 - 116) | 104 (94 - 126) | 103 (95 - 118) | 0.001 | <0.001 | <0.001 |
| HbA1c (%) | 5.4 (5.1 - 6.0) | 5.7 (5.3 - 6.6) | 5.9 (5.4 - 6.8) | 6.0 (5.6 - 6.8) | <0.001 | <0.001 | <0.001 |
| Plasma renin activity, ng/mL/hr | 0.7 (0.4 - 1.6) | 0.6 (0.3 - 1.2) | 0.8 (0.4 - 2.1) | 0.8 (0.4 -1.7) | 0.234 | 0.222 | 0.595 |
| Plasma aldosterone concentration, pg/mL | 127 (74 - 198) | 121 (78 - 187) | 125 (80 - 195) | 147 (74 - 215) | 0.812 | 0.662 | 0.718 |
| Comorbidities |  |  |  |  |  |  |  |
| Diabetes, n (%) | 47 (27.6) | 84 (39.3) | 62 (44.9) | 33 (45.8) | 0.023 | 0.002 | 0.009 |
| Dyslipidemia, n (%) | 97 (57.1) | 134 (62.6) | 96 (70.0) | 56 (77.8) | 0.317 | 0.033 | 0.004 |
| Anti-hypertensive drugs |  |  |  |  |  |  |  |
| Calcium-channel blocker, n (%) | 104 (68.9) | 129 (67.2) | 85 (69.1) | 52 (78.8) | 0.829 | 1.000 | 0.183 |
| α blocker, n (%) | 10 (6.6) | 14 (7.3) | 8 (6.5) | 11 (16.7) | 0.978 | 1.000 | 0.040 |
| Diuretic agent, n (%) | 8 (5.3) | 23 (12.0) | 14 (11.4) | 7 (10.6) | 0.051 | 0.105 | 0.260 |
| Sleep apnea |  |  |  |  |  |  |  |
| AHI | 2.4 (1.1 - 3.6) | 9.0 (7.1 - 11.5) | 20.1 (16.7 - 24.2) | 39.9 (33.0 - 45.6) | <0.001 | <0.001 | <0.001 |
| PA, n (%) | 45 (26.5) | 69 (32.2) | 34 (24.6) | 28 (38.9) | 0.264 | 0.814 | 0.077 |

Data are presented as the mean ± standard deviation or median (25th-75th percentile) for continuous variables, or number (%) for categorical variables. P values based on the two-sample Welch’s t-test or Mann-Whitney U-test are shown for comparisons of location parameters, such as mean or median of two groups, or P values based on the Fisher’s exact test for comparisons of percentage between groups. PA, primary aldosteronism; EH, essential hypertension; AHI, apnea-hypopnea Index; BP, blood pressure; LDL, low density lipoprotein; HDL, high density lipoprotein; eGFR, estimated glomerular filtration rate

### Relationship of PA with degree of OSA

Results of multivariable logistic analysis performed to examine the associations of PA with each level of OSA as compared to non-PA patients are shown in **Table 3**. This analysis was fully adjusted for clinical factors, including potential causal factors for sleep apnea, i.e., age, male gender, BMI, systolic BP, HbA1c, and renal function. PA was shown to have a significant association with only severe OSA. These results suggest that patients with PA are more likely than hypertensive patients without PA to be complicated by severe sleep apnea under adjustment of the potential causal factors for sleep apnea.

**Table 3.** Multivariable logistic regression analysis of factors associated with degree of sleep apnea

| Variables | Mild (AHI 5 to <15) |  | Moderate (AHI 15 to <30) |  | Severe (AHI ≥30) |  |
| --- | --- | --- | --- | --- | --- | --- |
|  | OR (95%CI) | P | OR (95%CI) | P | OR (95%CI) | P |
| Age, years (per 5) | 1.02 (0.95 - 1.11) | 0.560 | 1.18 (1.07 - 1.30) | 0.001 | 1.13 (1.00 - 1.05) | 0.084 |
| Male | 1.16 (0.82 - 1.63) | 0.410 | 1.29 (0.87 - 1.91) | 0.212 | 1.56 (0.88 - 2.78) | 0.130 |
| Body mass index, kg/m<sup>2</sup> (per 5) | 0.94 (0.78 - 1.13) | 0.492 | 1.35 (1.1 - 1.65) | 0.004 | 2.78 (2.08 – 3.73) | <0.001 |
| Systolic BP, mmHg (per 10) | 1.03 (0.92 - 1.16) | 0.615 | 1.04 (0.91 - 1.19) | 0.566 | 1.11 (0.92 – 1.34) | 0.275 |
| HbA1c (%) | 1.05 (0.92 - 1.20) | 0.485 | 1.08 (0.93 - 1.25) | 0.335 | 1.16 (0.94 - 1.43) | 0.174 |
| eGFR, mL/min/1.73 m<sup>2</sup> (per 5) | 1.00 (0.97 - 1.05) | 0.708 | 1.00 (0.95 - 1.05) | 0.897 | 0.93 (0.86 – 1.00) | 0.045 |
| PA (vs. non-PA) | 1.28 (0.86 - 1.90) | 0.229 | 0.97 (0.60 - 1.57) | 0.905 | 2.08 (1.09 – 3.95) | 0.025 |

PA, primary aldosteronism; BP, blood pressure; eGFR, estimated glomerular filtration rate; OR, odds ratio; CI, confidence interval

### Effects of elevated renin activity on change of sleep apnea

Renin activity and AHI were reevaluated after a mean of 2.5 ± 1.6 years in 45 PA patients. Pearson’s correlation coefficient were used to evaluate the association between the logarithm of the renin activity at reevaluation and amount of change in AHI, as shown in **Figure 2**. This relationship was significantly negatively correlated (r = -0.364, p = 0.014).

**Figure 2.**
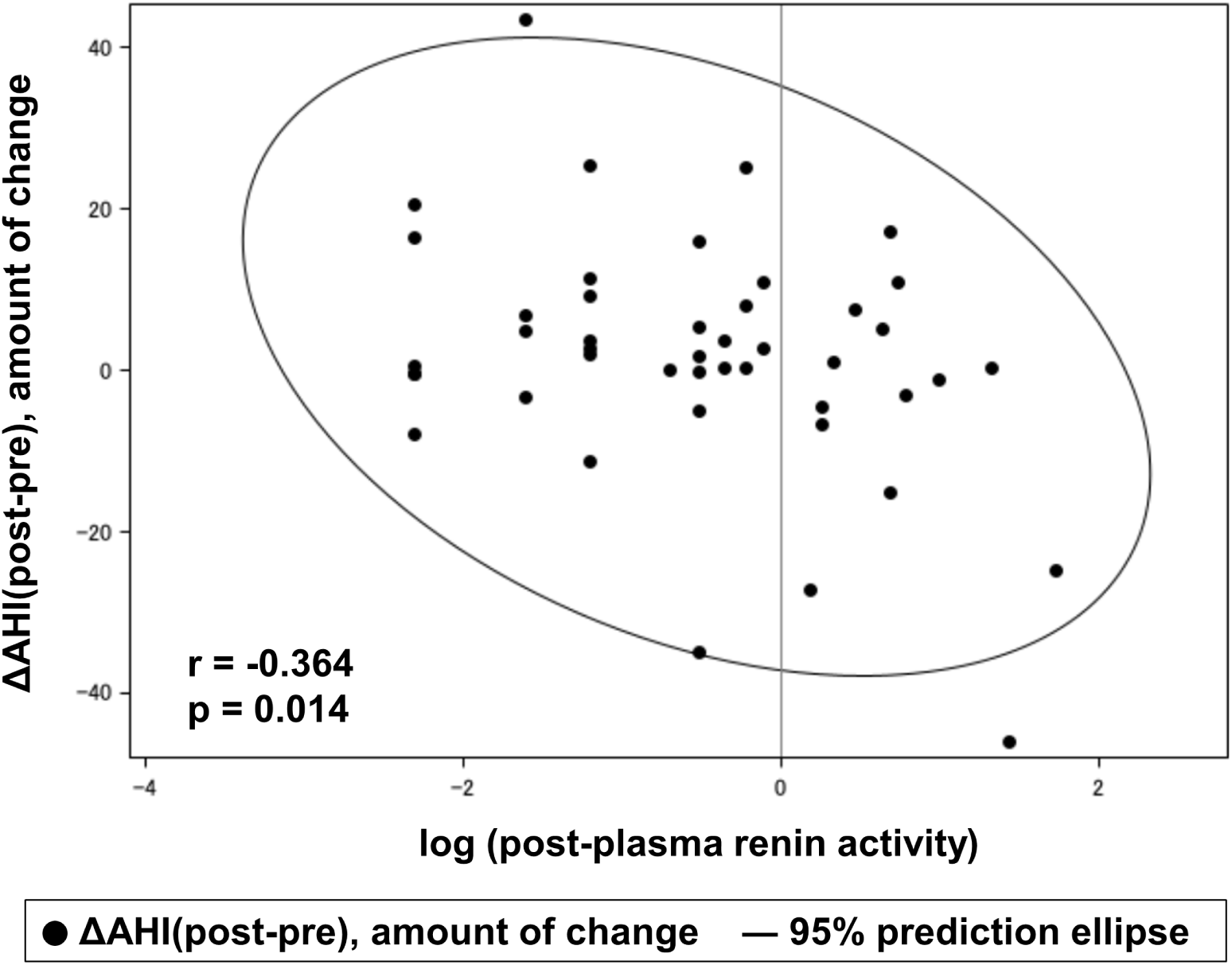
Association between renin activity following treatment and amount of changes in AHI. Correlation coefficient and p values for association between renin activity following treatment and amount of change in AHI were calculated using a Pearson’s correlation coefficient.

## Discussion

This is the first study to assess cross-sectional associations among PA patients with various degrees of OSA. Moreover, the effects of remaining suppressed renin activity on sleep apnea in patients with PA were prospectively examined. PA was found to be strongly associated with severe sleep apnea under adjustment of the potential causal factors for sleep apnea. Additionally, they show that elevated renin activity can potentially prevent the progression of sleep apnea in patients with PA.

Epidemiological studies have found that PA is a disease that can cause sleep apnea. Among patients with resistant hypertension, the degree of OSA has been reported to be higher in those with hyperaldosteronism ^14^. The present results show an association of PA with severe sleep apnea under adjustment of the potential causal factors for sleep apnea. Previously, it was suggested that excessive aldosterone secretion causes inappropriate sodium and fluid retention ^6^. Furthermore, mechanisms related to fluid movement from the lower extremities to the neck caused by edema of the upper airway may contribute to the etiology of OSA ^15^. Another study noted that patients with OSA who underwent diuretic treatment showed significantly improved upper airway caliber and OSA, further implicating pharyngeal edema as a cause of upper airway collapse during sleep ^16^. Moreover, a volumetric analysis technique used with magnetic resonance imaging demonstrated that volume of the tongue and lateral walls were independent factors related to increased risk of OSA ^17^. Impairment of ventilatory-control mechanisms can also contribute to modulation of pharyngeal collapsibility during sleep, thus suggesting a role for central nervous system input, which may be influenced by humoral factors ^18–20^. Furthermore, basic research results have indicated that aldosterone has a central role in increased brain renin-angiotensin activity, oxidative stress, and sympathetic drive ^21^. Therefore, in addition to pharyngeal edema caused by inappropriate sodium and fluid retention related to hyperaldosteronism, disruptions in normal central respiratory mechanisms by aldosterone-induced activation of central receptors may potentially contribute to the association between PA and sleep apnea severity.

Aldosterone receptor antagonists are used as therapeutic agents in patients with PA. Such medication has also been indicated as effective for not only improving hypertension but also preventing complications, including cardiovascular events. Spironolactone treatment for patients with resistant hypertension has been reported to significantly decrease AHI level ^22^. As for pharmacological treatment for PA, elevated renin activity has been shown useful to prevent development of cardiovascular events, cardiovascular death, and renal dysfunction, thus elevated renin activity is considered to be a therapeutic goal. A possible mechanism underlying the association between elevated renin activity and sleep apnea following therapeutic intervention may be inhibition of mineralocorticoid receptor activation by aldosterone antagonists or an adrenalectomy for aldosterone-producing adenoma treatment ^23^. Elevated renin activity is used as an indicator of suppressed mineralocorticoid receptor activation. Results of the present study indicate that elevated renin activity leads to prevent progress of sleep apnea.

This study has several limitations. First, the findings may not be generalizable to all patients with different blood pressure status. Second, the present cohort did not take into account the effect of change in body weight, as that was not possible to track. Nevertheless, the present findings are considered to provide important initial information for clarification of the association of PA with sleep apnea. Moreover, there was a significant negative correlation between the renin activity after treatment and change in AHI before and after treatment. The results suggest that the increase in renin activity reflects suppression of aldosterone action, which may lead to improvement of sleep apnea via improvement of pharyngeal edema.

## Conclusion

As compared to hypertensive patients without PA, patients with PA are more likely to have severe sleep apnea. Furthermore, elevated renin activity may contribute to improvement of sleep apnea in patients with PA.

## Perspectives

PA appears to play a fundamental role in development of sleep apnea. The elevated renin activity is used as an indicator of the effects of PA treatment goals as well as need for prevention of various complications. The present study investigated the association of PA with degree of OSA as compared to non-PA patients with hypertension and is the first to report a relationship between elevated renin activity and changes in sleep apnea in PA patients. The results provide important initial insight regarding the association of PA with degree of sleep apnea as well as for determining whether elevated renin activity has effects on sleep apnea in patients with PA.

## Data Availability

The datasets generated during and/or analyzed for the current study are not publicly available, though can be provided by the corresponding author upon reasonable request.

## Acknowledgements

The authors are grateful for the excellent technical assistance provided by Tomoe Ushitani and Ai Matsumoto. We also wish to thank the other investigators and staff members, as well as the participants of the HSCAA study for their valuable contributions.

## Novelty and Relevance

### What is new?

This study sheds light on the currently incompletely understood association between PA and degree of OSA in hypertensive patients and effect of elevated renin activity for change in sleep apnea in patients with PA. An increased risk of severe OSA is revealed in patients with PA. Furthermore, it indicated that elevated renin activity following treatment can possible to prevent sleep apnea progression.

### What Is Relevant?

Our results provide important initial findings to unveil the pathophysiological factors for PA and OSA.

### Clinical/Pathophysiological Implications?

PA is an important risk factor for severe OSA in hypertensive patients, and elevated renin activity following treatment is negatively correlated with progression of sleep apnea. These knowledge could improve the prediction and progression of sleep apnea in patients with PA.

